# A Multi-Site Analysis of the Prevalence of Food Security in the United States, before and during the COVID-19 Pandemic

**DOI:** 10.1101/2021.07.23.21260280

**Authors:** Meredith T. Niles, Alyssa W Beavers, Lauren A. Clay, Marcelle M. Dougan, Giselle A. Pignotti, Stephanie Rogus, Mateja R. Savoie-Roskos, Rachel E. Schattman, Rachel M. Zack, Francesco Acciai, Deanne Allegro, Emily H. Belarmino, Farryl Bertmann, Erin Biehl, Nick Birk, Jessica Bishop-Royse, Christine Bozlak, Brianna Bradley, Barrett P. Brenton, James Buszkiewicz, Brittney N. Cavaliere, Young Cho, Eric M. Clark, Kathryn Coakley, Jeanne Coffin-Schmitt, Sarah M. Collier, Casey Coombs, Anne Dressel, Adam Drewnowski, Tom Evans, Beth J Feingold, Lauren Fiechtner, Kathryn J. Fiorella, Katie Funderburk, Preety Gadhoke, Diana Gonzales-Pacheco, Amelia Greiner Safi, Sen Gu, Karla L. Hanson, Amy Harley, Kaitlyn Harper, Akiko S. Hosler, Alan Ismach, Anna Josephson, Linnea Laestadius, Heidi LeBlanc, Laura R. Lewis, Michelle M Litton, Katie S. Martin, Shadai Martin, Sarah Martinelli, John Mazzeo, Scott C. Merrill, Roni Neff, Esther Nguyen, Punam Ohri-Vachaspati, Abigail Orbe, Jennifer J. Otten, Sondra Parmer, Salome Pemberton, Zain Al Abdeen Qusair, Victoria Rivkina, Joelle Robinson, Chelsea M. Rose, Saloumeh Sadeghzadeh, Brinda Sivaramakrishnan, Mariana Torres Arroyo, McKenna Voorhees, Kathryn Yerxa

## Abstract

**Background:** The COVID-19 pandemic profoundly affected food systems including food security. Understanding how the COVID-19 pandemic impacted food security is important to provide support, and identify long-term impacts and needs.

**Objective:** Our team- the National Food Access and COVID research Team (NFACT) was formed to assess food security over different U.S. study sites throughout the pandemic, using common instruments and measurements. Here we present results from 18 study sites across 15 states and nationally over the first year of the COVID-19 pandemic.

**Methods:** A validated survey instrument was developed and implemented in whole or part across the sites throughout the first year of the pandemic, representing 22 separate surveys. Sampling methods for each study site were convenience, representative, or high-risk targeted. Food security was measured using the USDA six-item module. Food security prevalence was analyzed using analysis of variance by sampling method to statistically significant differences.

**Results:** In total, more than 27,000 people responded to the surveys. We find higher prevalence of food insecurity (low or very low food security) since the COVID-19 pandemic, as compared to before the pandemic. In nearly all study sites, we find higher prevalence of food insecurity among Black, Indigenous, and People of Color (BIPOC), households with children, and those with job disruptions. We also demonstrate lingering food insecurity, with high or increased prevalence over time in sites with repeat surveys. We find no statistically significant differences between convenience and representative surveys, but statistically higher prevalence of food insecurity among high-risk compared to convenience surveys.

**Conclusions:** This comprehensive multi-study site effort demonstrates higher prevalence of food insecurity since the beginning of the COVID-19 pandemic, which in multiple survey sites continues throughout the first year of the pandemic. These impacts were prevalent for certain demographic groups, and most pronounced for surveys targeting high-risk populations.

## Introduction

The coronavirus disease 2019 (COVID-19) was declared a pandemic by the World Health Organization in March 2020 (1), with widespread impact across the United States (U.S.) and globally. As of April 12, 2021, the U.S. had over 20% of confirmed cases and about 19% of the COVID-19-related deaths globally (2). Furthermore, COVID-19 was the third leading cause of death in the U.S. in 2020 (3).

The pandemic caused major disruptions to the U.S. economy, food system, and overall health and wellbeing of Americans. The unemployment rate in the U.S. reached an unprecedented high of 14.8% in April 2020 (4), with job disruptions concentrated in low-paying jobs, disproportionately affecting Black, Indigenous, and People of Color (BIPOC) (5). Although the unemployment rate declined to 6.7% in December 2020, the economic effects of the pandemic are likely to persist for years, consistent with the Great Recession of 2008 (6). The need to social distance and quarantine to contain disease spread led to stockpiling, placing a strain on the food supply chain, which was unable to adequately respond to the pandemic, resulting in food access concerns for many Americans (7). This, in combination with widespread disruption in employment, increased food-related hardship for many Americans, particularly those most vulnerable to economic disruption (8).

Disasters, like hurricanes, and public health emergencies like the COVID-19 pandemic disrupt built and social environments, and their impacts persist long after they occur (9-11). Disasters tend to impact housing stability, household composition, and financial obligations, which can limit resources for food and lead to food-related hardship (12). Groups most vulnerable to disasters were disproportionately affected during the pandemic, including low-income households, single-headed households with children, adults living alone, and Black- and Hispanic-headed households (13-15). The COVID-19 pandemic magnified the health disparities that exist among low-income households, who were already more likely to struggle to meet basic needs (15).

Food insecurity, or the inability to consistently obtain enough, desirable, varied, and nutritious foods (16), is heightened during disasters and emergencies (17, 18). Emergency nutrition response aims to assist affected individuals; however coordinating enough high-quality food remains a challenge in a post-disaster setting (19, 20). Quickly assessing food insecurity to inform pandemic relief efforts was a challenging task; for instance, the national food insecurity statistics for 2020 from the U.S. Census Bureau, measured using the USDA’s Household Food Security Survey Module (HFSS), will not be released until September 2021 (21). As a result, agencies, organizations, and researchers deployed surveys and produced estimates to determine the impact of the pandemic on food insecurity. For example, the U.S. Census Bureau released the Household Pulse Survey that captures food insufficiency and Feeding America released projected food insecurity prevalence for 2020 and 2021 based on changes in unemployment and poverty (21-23). Nationally representative surveys found that food insecurity drastically increased at the onset of the COVID-19 pandemic, from 11% in 2018 up to 38% in March 2020 (24). This is especially high considering the impact of economic downturn during the Great Recession of 2008 when food insecurity peaked at much lower 15% in 2011 (13, 25). In addition, households that were food insecure prior to the pandemic were more likely to have their situations exacerbated due to less job flexibility, higher risk of job loss/furlough, and fewer resources/support to allow for complying with social distancing recommendations (15). Although the early months of the pandemic may have been the peak of food insecurity and insufficiency, higher than usual rates have persisted as the COVID-19 pandemic continues (26).

Despite several early surveys assessing food insecurity during the COVID-19 pandemic, and continued efforts to measure food insufficiency through the Census Household Pulse Survey, there have been few collaborative efforts to monitor and measure food insecurity across diverse geographic and social contexts, and to compare data. In May 2020, a national collaboration of researchers - The National Food Access and COVID Research Team (NFACT) - was formed to examine COVID-19 impacts on food access, food insecurity, and the overall food system. This study reports the findings of this collaborative effort, with data from 18 study sites including a nationally representative sample, to better understand food insecurity over diverse regions and timeframes. The study examined overall levels of food insecurity, as well as food insecurity among households with children, households that experienced job disruption, and participants identifying as BIPOC. We further assessed how different survey implementation methods associate with different levels of food insecurity, and report results from multiple time points within the same study site, based on data availability.

## Methods

### Survey Development

A survey instrument, known as the NFACT Survey Version 1.0 (27) was developed in March 2020. This survey was developed in consultation with key stakeholders in the state of Vermont, where it was first implemented, and drew from the existing literature on food security and food access. Where possible, validated questions and instruments were used. The survey was piloted in Vermont, with 25 adult residents in late March, and validation methods (e.g. Cronbach alpha, factor analysis) were used to test the internal validity of questions with key constructs (alpha > 0.70) (28). A second version of the survey was released in May 2020 to reflect changes in the COVID-19 context (29) and include new questions. The surveys included questions on food access, food security, food purchasing, food assistance program participation, dietary intake, perceptions of COVID-19, and individual and household sociodemographic characteristics. The questions utilized in this study were included in both surveys and across study sites.

### NFACT Study Sites and Data Collection

NFACT represents 18 study sites across 15 states, as well as a national sample (Figure 1). NFACT study sites distributed the NFACT surveys (in whole or part) online pursuing one of three sampling strategies: 1) Convenience sampling in partnership with community organizations, stakeholders, social media, and/or news media, which are not representative of a state population (ten sites); 2) Quota sampling using survey panels administered by Qualtrics (Provo, UT), a survey research company, in which the quotas aimed to achieve state representation on some characteristics (e.g. race, ethnicity, income) (eight sites); or 3) Quota or convenience sampling in which certain high-risk populations (e.g. low-income, BIPOC, or Supplemental Nutrition Assistance Program (SNAP) participants) were targeted (six sites). In some cases where high-risk populations were targeted, these groups were oversampled to ensure adequate representation in the overall study sample. Table 1 provides specific details about the sampling strategies, target populations, representation of the data, and survey fielding dates. Potential participants under age 18 were excluded across all study sites. All study sites administered the survey in English; in Arizona, California-Bay Area, Maine, Massachusetts, Nationally, NY-Capital Region, New Mexico, and Utah, surveys were also administered in Spanish. IRB approval was obtained by each study site prior to commencing data collection.

**Figure 1.**
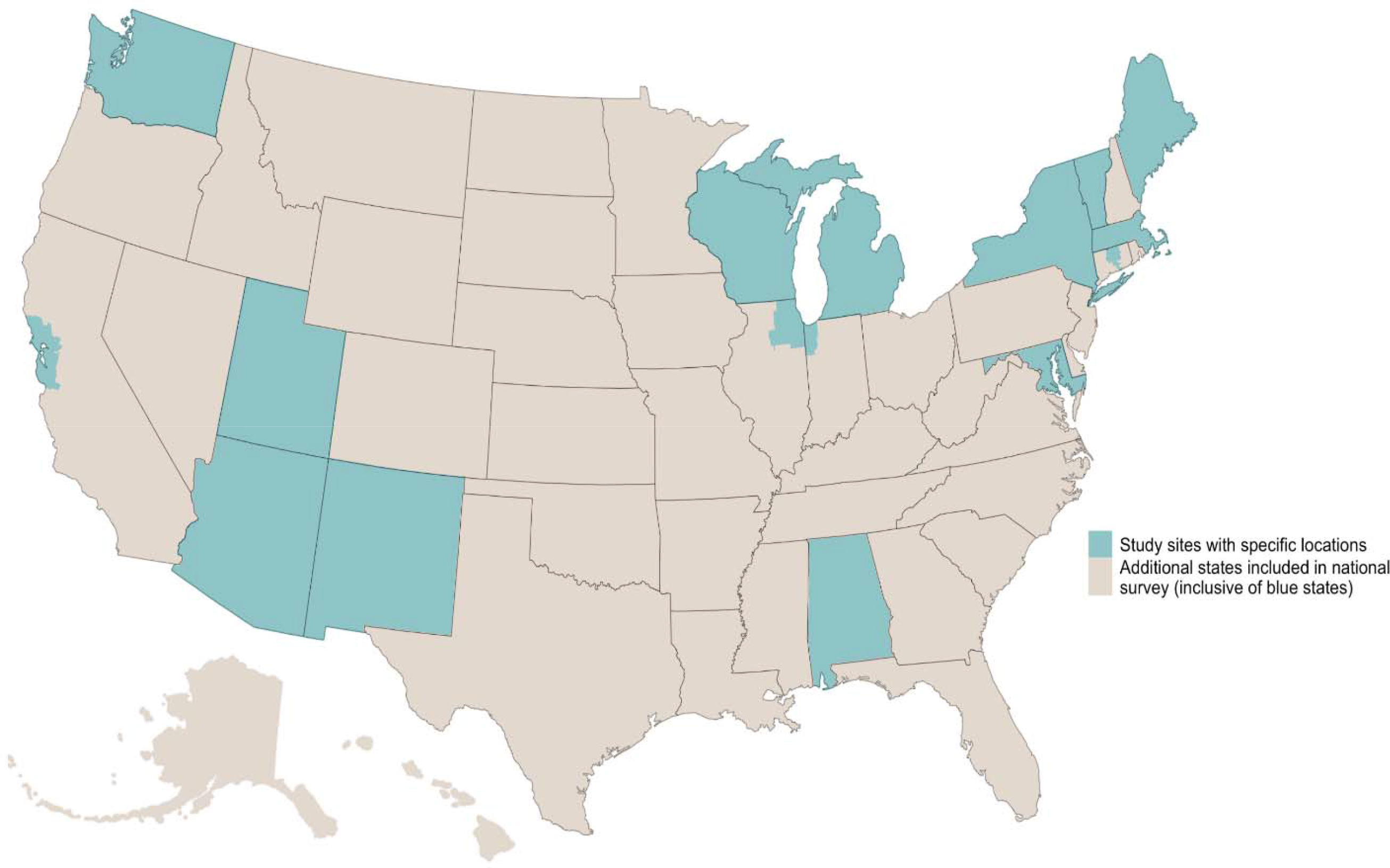
NFACT study sites. Blue states and regions represent sites in addition to the national sample strategy, which includes additional data from all states. Visual credit: Samuel F. Rosenblatt

**Table 1.**
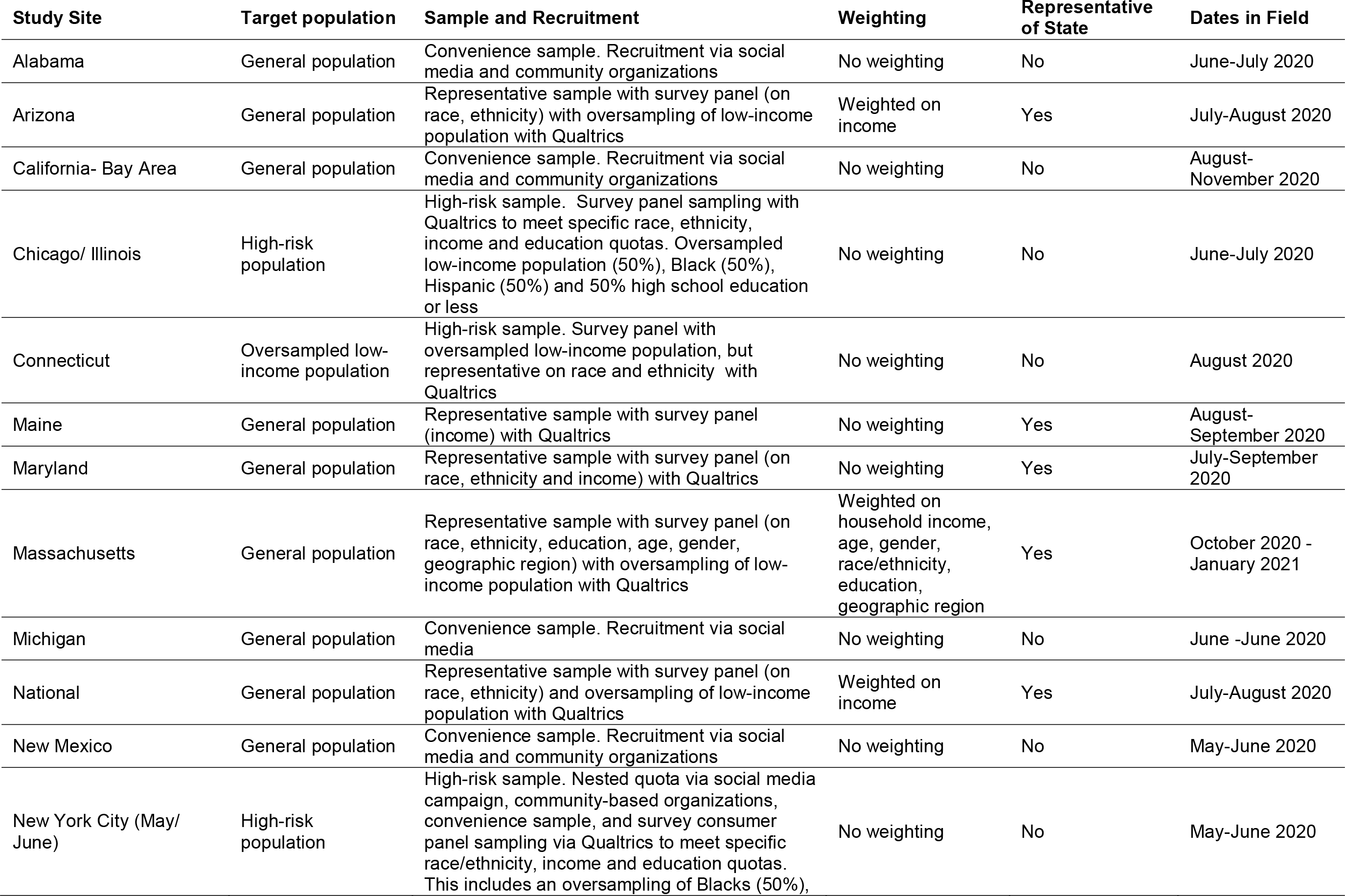
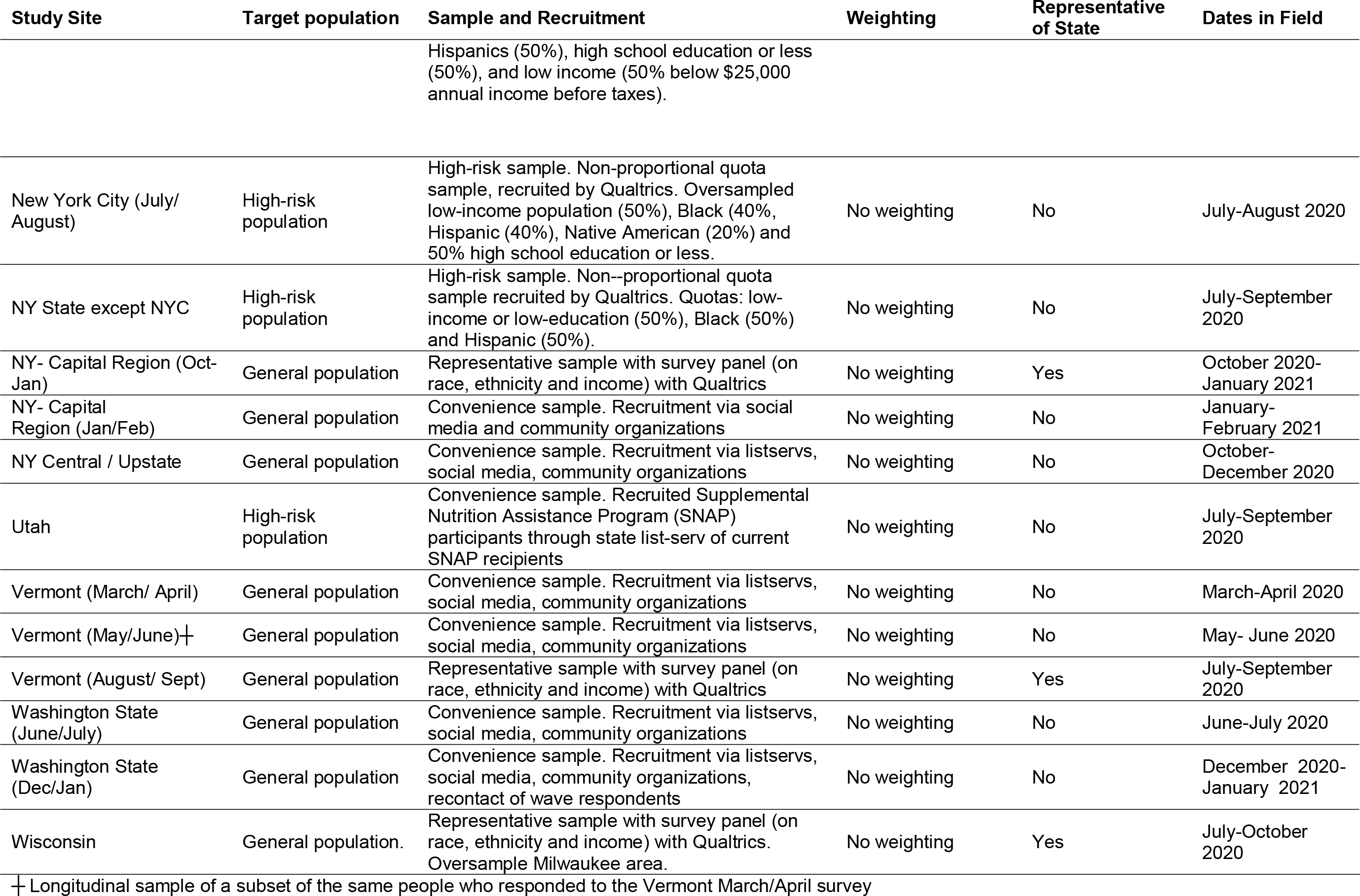
Study sites and relevant methods for each site.

### Measures

Food security was assessed using the United States Department of Agriculture (USDA) 6-item Short Form Food Security Survey Module (30) which is designed to identify households with food insecurity. In most sites, participants were asked to complete 6-items about the year before COVID-19 and since the COVID-19 pandemic began in March 2020, though a few sites only asked these questions since the COVID-19 pandemic began. In some more recent surveys (i.e. Massachusetts, NY-Central/Upstate and the second Washington survey) and in Michigan respondents answered questions about food security in the past 30 days, which is validated through the USDA module. Following standard USDA scoring, a score of 2-6 was categorized as food insecure (30). It is important to note that the pre-pandemic food security responses were retrospective, and were answered at the same time as the questions about current food security. Households with children were determined with a question about household composition by age. Households with any members ages 0-17 years were classified as a household with children. Job disruption was assessed by asking participants if their household experienced a job disruption since the start of the pandemic, including job loss, furlough, or loss of hours/income reduction, categories which were not mutually exclusive. Participants indicating any negative job impact were categorized as experiencing a job disruption. BIPOC classification was determined based on survey questions about race and ethnicity. Participants indicating any race or ethnicity besides non-Hispanic White (NHW) were classified as BIPOC. Participants indicating NHW were classified as such and Hispanic of any race were classified as Hispanic.

### Data aggregation and analysis

Food insecurity prevalence (overall and for specific populations of interest) by study site and survey were aggregated into a single dataset for analysis in Stata 16.0 (31). While we primarily report descriptive statistics of the results across the multiple sites, we also used analysis of variance (ANOVA) with Scheffe multiple comparison tests (32) to assess whether there are statistically significant differences in prevalence of food insecurity (overall and for key sub-populations) between surveys based on the three different sampling techniques (i.e. convenience, representative, and high-risk). We report p values <0.05 as statistically significant in the results.

## Results

### Respondent Characteristics

The sample included 27,168 adults from across the U.S. with data on food insecurity. The racial and ethnic make-up of the sample overall was 70.0% NHW, and 28.6% BIPOC, with 1.4% of respondents not identifying race or ethnicity. Among BIPOC respondents, 8.0% identified as non-Hispanic Black, 11.9% as Hispanic, and 8.1% other races or multiracial (Table 2). Given the diversity of NFACT study sites, including their sample size and demographic make-up, the number of respondents with demographic characteristics or life experiences (e.g. job disruption or children in the household) varied across study sites. There was a large variation in the proportion of BIPOC respondents across study sites, because of differences in population composition, but also because some study sites oversampled BIPOC respondents. Slightly over 40% of respondents (40.6%) had children in the household, ranging from 19.2% of households in Maine to 85.6% of households in the California, Bay Area. Among all respondents, 35.3% had experienced some type of job disruption since the COVID-19 pandemic began, ranging from a low of 10.8% of respondents in a second Washington State survey in early 2021, to 76.5% of respondents in a NY Capital Region survey in January and February 2021. Among representative samples, the range varied from 37.6% in the national sample to 61.3% in the NY-Capital region in October-January.

**Table 2.** Total number of respondents and sub-population characteristics by study site.

| Study Site | Total Respondents<sup>1</sup> | With Children | Job Disruption/<br>(Reduced Income) | BIPOC<sup>2</sup> | NHW<sup>3</sup> | NHB<sup>4</sup> | Hispanic | Other or Multiple Races |
| --- | --- | --- | --- | --- | --- | --- | --- | --- |
| Alabama | 1247 | 541 | 546 | 226 | 1061 | 142 | 27 | 86 |
| Arizona | 576 | 189 | 221 | 268 | 352 | 32 | 194 | 42 |
| California- Bay Area | 724 | 620 | 321 | 232 | 223 | 6 | 122 | 49 |
| Chicago/ Illinois | 680 | 379 | 314 | 498 | 169 | 215 | 258 | 103 |
| Connecticut | 512 | 199 | 286 | 158 | 354 | 56 | 73 | 54 |
| Maine | 504 | 97 | 193 | 42 | 477 | 9 | 8 | 8 |
| Maryland | 903 | 330 | 368 | 427 | 555 | 239 | 91 | 97 |
| Massachusetts | 2939 | 1098 | 1467 | 748 | 2191 | 202 | 292 | 254 |
| Michigan | 484 | 237 | 279 | 64 | 418 | 25 | 18 | 21 |
| National | 1510 | 515 | 568 | 585 | 925 | 212 | 255 | 118 |
| New Mexico | 1415 | 406 | 261 | 494 | 843 | 15 | 362 | 117 |
| New York City (May/<br>June) | 1,165 | 599 | 494 | 876 | 289 | 252 | 496 | 128 |
| New York City<br>(July/August) | 525 | 317 | 285 | 484 | 41 | 154 | 123 | 102 |
| NY State | 494 | 207 | 189 | 494 | n/a | 260 | 234 |  |
| NY -Capital Region<br>(Oct-Jan) | 479 | 167 | 294 | 156 | 353 | 43 | 42 | 71 |
| NY-Capital Region<br>(Jan-Feb) | 427 | 283 | 327 | 137 | 317 | 62 | 56 | 19 |
| NY- Central/Upstate | 434 | 120 | 144 | 30 | 380 | 2 | 10 | 22 |
| Utah | 644 | 219 | 277 | 102 | 392 | 12 | 61 | 56 |
| Vermont (March<br>/April) | 3016 | 913 | 1103 | 150 | 2603 | 5 | 45 | 104 |
| Vermont (May/<br>June) | 1212 | 383 | 294 | 57 | 1137 | 3 | 19 | 37 |
| Vermont (August/<br>Sept) | 578 | 178 | 270 | 49 | 551 | 6 | 17 | 26 |
| Washington State<br>(June/July) | 2514 | 1095 | 636 | 592 | 1910 | 93 | 210 | 289 |
| Washington State<br>(Dec/Jan) | 3169 | 1541 | 343 | 737 | 2647 | 98 | 283 | 356 |
| Wisconsin | 1017 | 393 | 430 | 181 | 836 | 58 | 80 | 43 |
| TOTAL | 27168 | 11026 | 9589 | 7787 | 19024 | 2195 | 3235 | 2202 |
| % of total |  | 40.6% | 35.3% | 28.7% | 70.0% | 8.1% | 11.9% | 8.1% |

1 Indicates number of total respondents with food security data
2 Black, Indigenous, People of Color respondents. Number includes anyone identifying as other than non-Hispanic white.
3 Non-Hispanic White (NHW)
4 Non-Hispanic Black (NHB)

### Overall Prevalence of Food Insecurity

We found higher levels of food insecurity reported since the COVID-19 pandemic began, as compared to reported for the pre-COVID-19 pandemic period. This finding was consistent in all 20 sites that asked about food insecurity both before and during the COVID-19 pandemic (Figure 2), with the exception of the New Mexico site (where no change was found). The prevalence of food insecurity across study sites during the COVID-19 pandemic ranged from 10.8% in a Central/Upstate New York convenience survey from October-December 2020 (which asked about the last 30 days), to 73.9% in a New York City high-risk survey in July/August 2020 which oversampled BIPOC, low-income respondents. Among states that represented state characteristics, food insecurity prevalence ranged from 28.8% in Maryland to 36.2% in Wisconsin since the start of the COVID-19 pandemic. In sites that gathered data on the time periods both before and during the COVID-19 pandemic, the rate of increase ranged from 0% in New Mexico to a 65% increase among respondents in the California Bay Area. We found that both convenience and representative samples had significantly lower prevalence of food insecurity both before and since the COVID-19 pandemic, as compared to surveys targeting high-risk populations, though the percent change did not significantly differ across survey sample type (Table 3).

**Figure 2.**
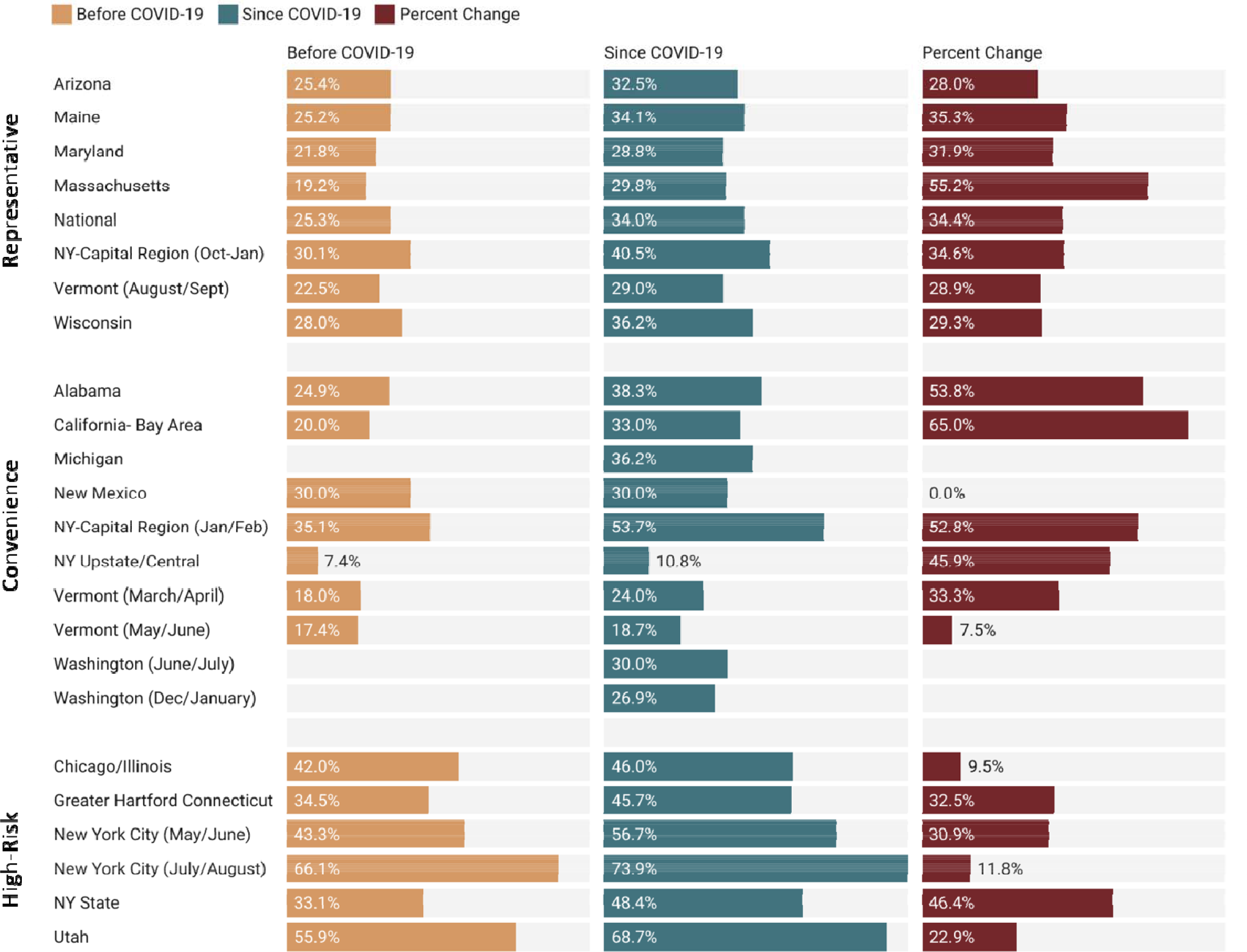
Overall prevalence of food insecurity across NFACT surveys and study sites.

**Table 3.** Overall prevalence of food insecurity across different measures and time periods by survey type. P values that are statistically significant (p<0.05) are lightly shaded for emphasis, and were obtained through ANOVAs with Scheffe multiple comparisons.

| Prevalence of Food Insecurity | Timeframe | Survey Type |  |  | P Values |  |  |
| --- | --- | --- | --- | --- | --- | --- | --- |
|  |  | Convenience | State Representative | High Risk | Convenience-High Risk | Convenience-Representative | Representative-High Risk |
| Overall Food Insecurity | Before COVID-19 | 21.8% | 23.9% | 43.6% | 0.002 | 0.918 | 0.004 |
|  | Since COVID-19 | 30.2% | 32.1% | 54.3% | 0.000 | 0.933 | 0.002 |
|  | Percent Change | 36.9% | 34.7% | 26.9% | 0.561 | 0.971 | 0.701 |
| BIPOC Food Insecurity | Before COVID-19 | 29.5% | 32.1% | 37.3% | 0.211 | 0.818 | 0.497 |
|  | Since COVID-19 | 40.2% | 40.6% | 55.1% | 0.048 | 0.999 | 0.078 |
|  | Percent Change | 32.0% | 29.5% | 36.0% | 0.892 | 0.957 | 0.743 |
| Households with Children Food Insecurity | Before COVID-19 | 30.1% | 37.2% | 44.1% | 0.042 | 0.389 | 0.424 |
|  | Since COVID-19 | 39.0% | 49.2% | 57.6% | 0.003 | 0.117 | 0.272 |
|  | Percent Change | 31.8% | 33.4% | 32.7% | 0.995 | 0.983 | 0.997 |
| Job Disruption Food Insecurity | Any Job Disruption | 43.5% | 50.1% | 64.8% | 0.003 | 0.489 | 0.058 |
|  | Job Loss | 51.3% | 60.8% | 72.1% | 0.003 | 0.216 | 0.168 |
|  | Furlough | 44.2% | 51.2% | 63.1% | 0.081 | 0.679 | 0.383 |

### Prevalence of Food Insecurity Among BIPOC Respondents

In all survey sites that collected data on food insecurity before and during the COVID-19 pandemic, we found that food insecurity increased for BIPOC respondents since the onset of the COVID-19 pandemic, with the exception of New Mexico. Furthermore, we found that the prevalence of food insecurity among BIPOC respondents during the COVID-19 pandemic was higher than the overall prevalence of food insecurity in the majority of study sites (Figure 3); however, it is worth noting that this was also true for pre-COVID-19 food insecurity. The highest percent increase in food insecurity was identified in the California Bay Area (54.2% increase in food insecurity among BIPOC respondents). However, the highest prevalence of food insecurity during the COVID-19 pandemic among BIPOC respondents was identified in the NY Capital Region (83.8%). We found the prevalence of BIPOC food insecurity during the COVID-19 pandemic was significantly different (p=0.048) for convenience (40.2%) versus high-risk (55.1%) survey types.

**Figure 3.**
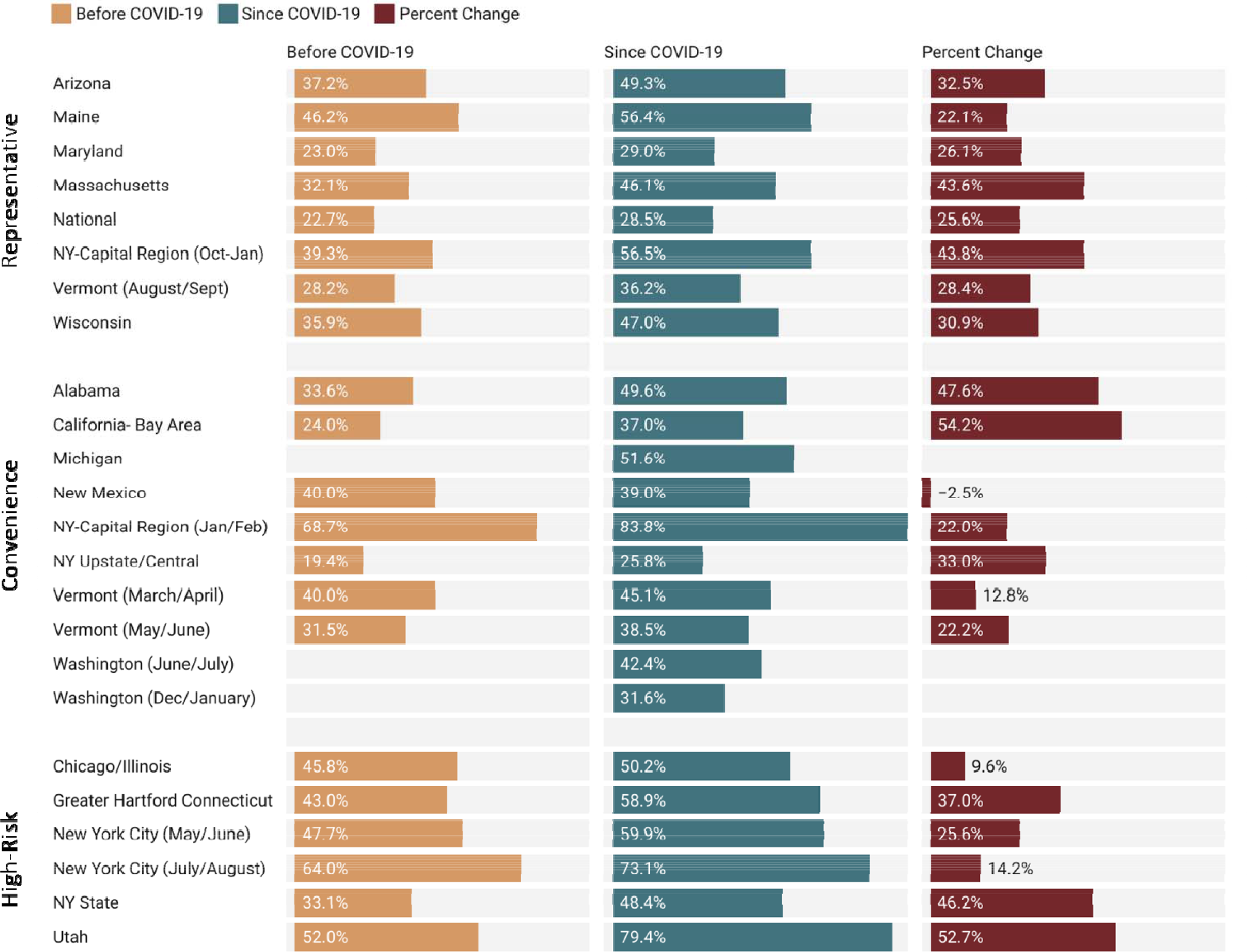
Prevalence of food insecurity before and during the COVID-19 pandemic, and the percent change, among BIPOC respondents, by study site.

Furthermore, we disaggregated race and ethnicity data when a particular survey had at least 30 respondents identifying within a specific race or ethnic group (Figure 4). This additional breakdown further highlights disparities in food insecurity across many study sites among BIPOC respondents, as compared to NHW respondents. For example, while the majority of surveys found the prevalence of food insecurity was higher for BIPOC respondents both before and during the COVID-19 pandemic, the opposite is true of NHW respondents (i.e. the majority of surveys found the prevalence of food insecurity among NHW respondents before and during the COVID-19 pandemic was lower than the site’s overall food insecurity).

**Figure 4.**
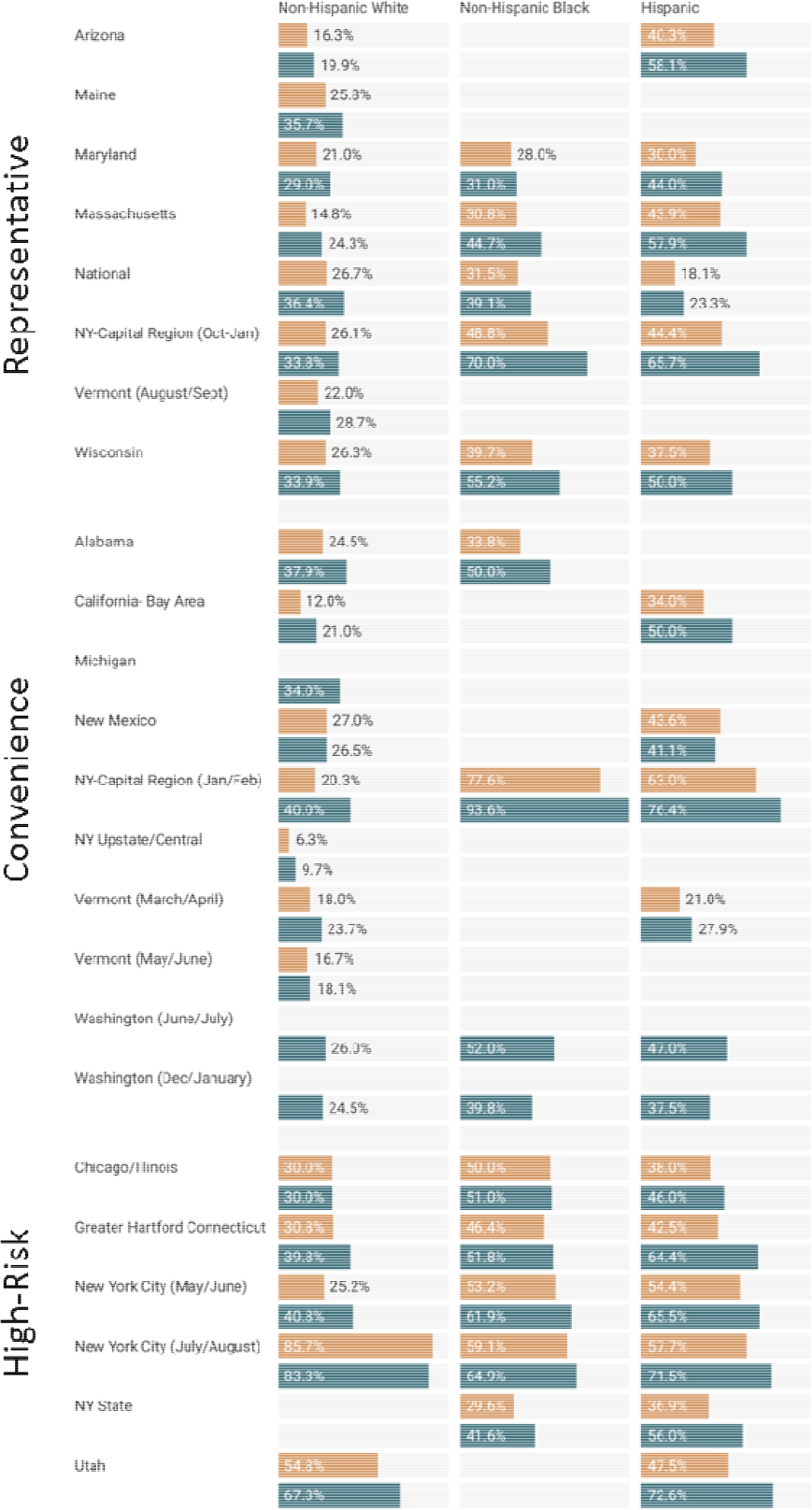
Prevalence of food insecurity before and during the COVID-19 pandemic, and the percent change, among different racial and ethnic groups, by study site. Disaggregated race and ethnicity food insecurity prevalence is only reported for sites where at least 30 respondents identified as a specific race or ethnic group.

### Prevalence of Food Insecurity Among Households with Children

In all but one survey (New Mexico) with data on food insecurity before and during the COVID-19 pandemic, food insecurity increased among households with children (Figure 5). The highest reported percent change was in Massachusetts (a 62.1% increase), while the overall highest prevalence during the COVID-19 pandemic was 69.3% food insecurity among households with children in a Utah survey focused on SNAP participants. In surveys representative of the state population, the prevalence of food insecurity among households with children ranged from 41.7% in Vermont in August/September 2020 to 56% in Arizona. Convenience surveys had statistically lower food insecurity prevalence as compared to high-risk survey populations both before the COVID-19 pandemic (p=0.042), and during the COVID-19 pandemic (p=0.003) (Table 3).

**Figure 5.**
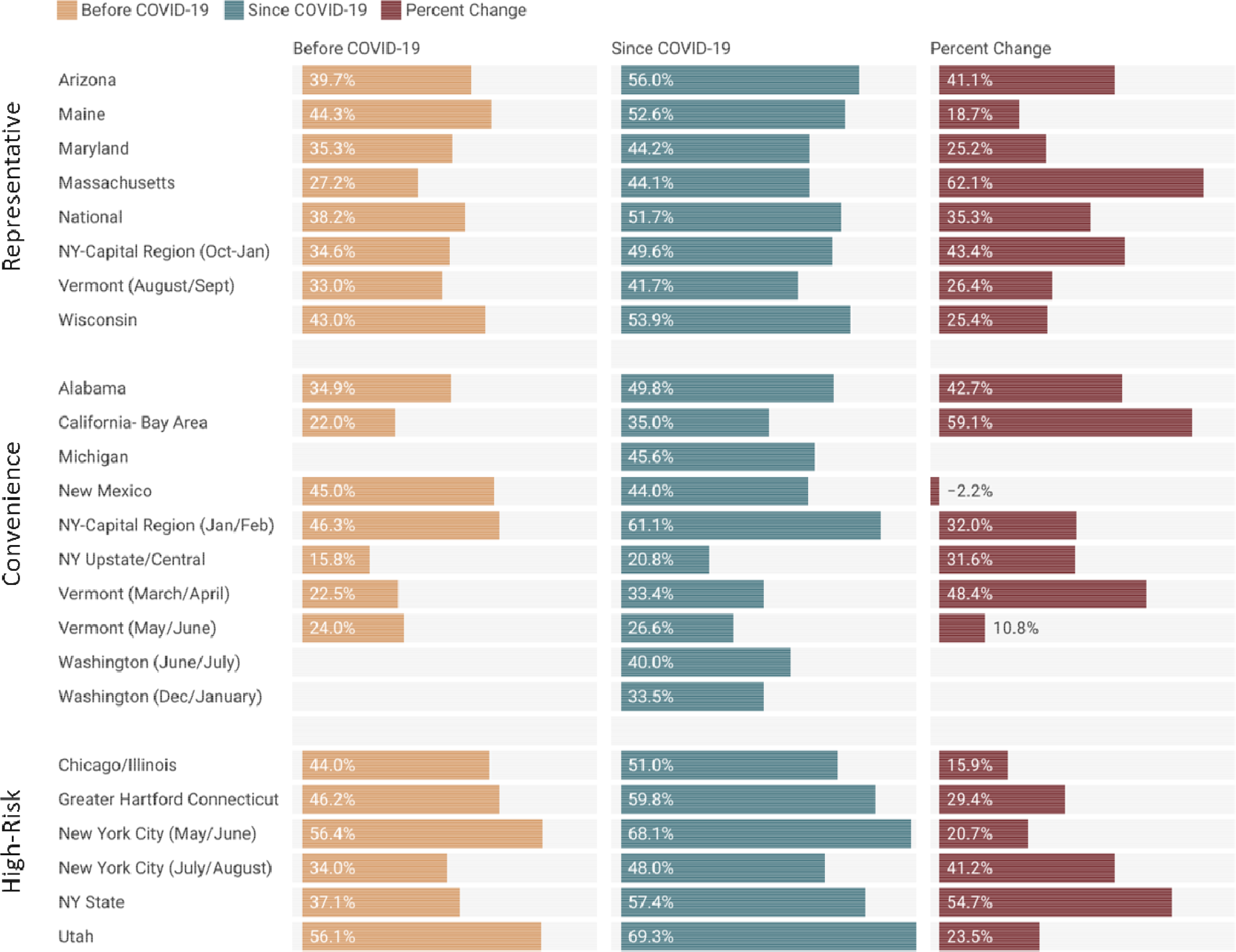
Prevalence of food insecurity before and during the COVID-19 pandemic among households with children in a study site, and the percent change.

### Prevalence of Food Insecurity Among Respondents Experiencing Job Disruption

Food insecurity during the COVID-19 pandemic was higher in all surveys and study sites among respondents facing a job disruption, as compared to the overall prevalence of food insecurity in those sites (Figure 6). The range of food insecurity among respondents with job disruptions ranged from 21.5% in Central/Upstate New York up to 77.2% in New York City among all surveys. Among surveys with state-wide representative samples on some characteristics, the prevalence of food insecurity for those with job disruptions ranged from 38.7% in Vermont in August/September 2020 to 59.8% in Wisconsin. Convenience surveys had statistically lower food insecurity prevalence as compared to high-risk survey populations for any job disruption (p=0.003), job loss (p=0.003), and reduction in hours (p=0.036) (Table 3).

**Figure 6.**
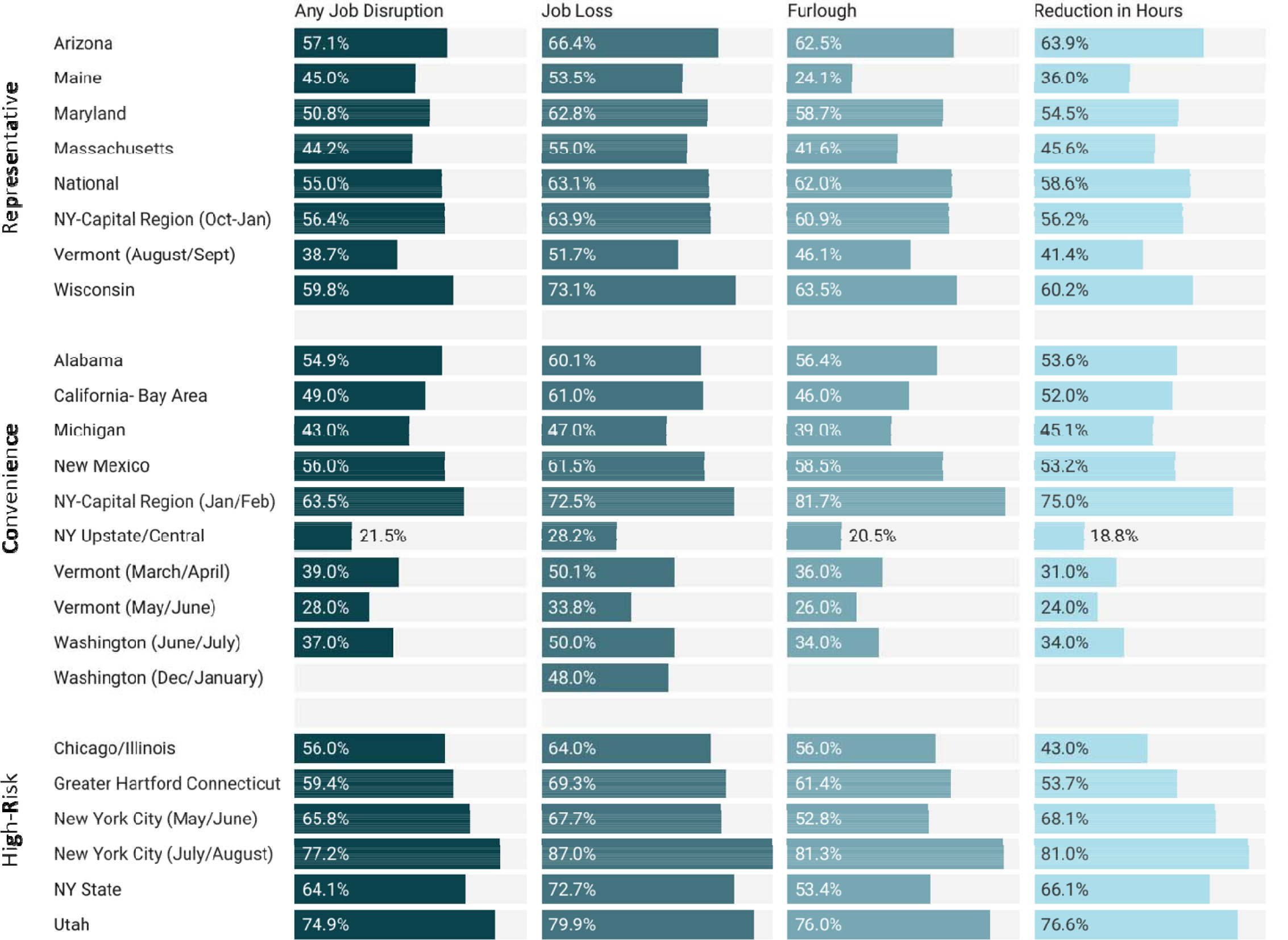
Prevalence of food insecurity since the COVID-19 pandemic among respondents with any job disruption, job loss, furlough, and/or reduction in hours, by study site.

## Discussion

In this study, food insecurity was assessed in multiple sites using a common measurement instrument. Key trends in food insecurity were highly consistent among research sites, albeit with some significant differences in magnitude depending on survey type. This study utilized three different sampling methods (representative, convenience, and targeted high-risk populations), allowing us to compare results between both study sites and sampling strategies. Notably, there were no statistically significant differences in our findings between convenience and representative samples, though high risk populations were consistently more likely to report food insecurity than those recruited through convenience samples. Nearly all study sites that assessed both current and pre-COVID-19 food insecurity found a higher prevalence of food insecurity during the COVID-19 pandemic as compared with before the COVID-19 pandemic.

Furthermore, the majority of surveys and sites found higher prevalence among BIPOC respondents as compared to the overall food insecurity prevalence and that of NHW respondents in the same area. All but one survey found higher prevalence of food insecurity for households with children during the COVID-19 pandemic as compared to the overall food insecurity prevalence in a given site, and all surveys found higher prevalence of food insecurity among respondents reporting job disruptions compared to those with no job disruptions. Importantly, among study sites that conducted repeated surveys, all found continued increase in food insecurity as the pandemic continued, demonstrating the ongoing and escalating effects of the COVID-19 pandemic. Below we further elaborate on three key findings, and discuss their implications for future programming and policy.

First, food insecurity increased across nearly all research sites between the pre- and during-pandemic periods. These results are consistent with several other national surveys examining the impact of COVID-19 on food insecurity. For example, data from the Census Household Pulse Survey and the COVID Impact Survey used probability sampling to obtain nationally representative samples. In the COVID Impact Survey, data collected in early April 2020 was extended using models to show that the overall prevalence of food insecurity was more than double the predicted rate (33). These same researchers found similar estimates of food insecurity increases using data from the Census Household Pulse Survey (34). The NFACT survey results support these findings across study sites, where direct data collection has occurred (as opposed to modelled results). Notably, NFACT sites that utilized a representative high-risk sampling approach were more likely than surveys using a convenience or representative sample to document higher prevalence of food insecurity since the onset of the COVID-19 pandemic. These results suggest that targeted oversampling of high-risk populations is likely to detect higher food insecurity outcomes, an important finding for future surveys and methodologies. Furthermore, when assessing overall food insecurity before or during the COVID-19 pandemic there were no statistically significant differences in food insecurity prevalence between convenience and representative sampling approaches. Among all survey approaches there were no significant differences in the percent change of prevalence of food insecurity, suggesting that the rate of change was fairly consistent across all survey types. These results provide important findings for researchers who must balance different priorities when determining a sampling approach in the future (e.g. cost, timeframe for data collection, ability to represent data at a state-level).

It should also be noted that our results show clear differences in food insecurity in different U.S. regions. These differences may be partially attributed to problems in the food supply-chain and community purchasing behavior (i.e. stockpiling), especially at the beginning of the pandemic (33). Another likely cause for variation is the inconsistent national approach to pandemic related restrictions such as stay-at-home orders, restrictions on businesses, and quarantine requirements. Variation in state response to the threat of rising food insecurity is best exemplified in state waivers authorized through SNAP and the Women, Infants and Children (WIC) program and administered through the USDA Food and Nutrition Services (FNS). Specifically, states had discretion about which benefits and waivers to request. While some states made repeated requests for a wide range of allowances authorized by Congress, others requested only a few (35, 36). It is likely that variation in states’ applications of extra benefits and temporary waivers influenced differences in food insecurity across our study sites. We suggest that future research examine the relative effects of extra benefits and waivers granted to states, and their influence on both programmatic enrollment and food security outcomes.

Second, our study found that some populations have experienced higher rates of food insecurity since the COVID-19 pandemic. Consistent with recent studies (33, 37), BIPOC populations reported higher rates of food insecurity than NHW respondents in nearly all NFACT study sites both before and since the onset of the COVID-19 pandemic. Moreover, the three sampling approaches used by NFACT sites found strikingly similar results. There was no statistical difference between sampling strategies with the exception of convenience and targeted high-risk approaches, specifically when addressing food insecurity among BIPOC respondents during the COVID-19 pandemic. Several other national surveys using professional survey platforms (Qualtrics and Turk Prime) have similarly found higher food insecurity rates among Black and Hispanic respondents compared with NHW respondents (15, 24). The only study to provide food insecurity data for Native American respondents found that this population also has a higher rate of food insecurity than NHW populations since the beginning of the pandemic (24). Our research and the work of others (38, 39) clearly shows that the short-term effects of the pandemic expose underlying racial and economic inequalities, but also highlights that BIPOC respondents faced higher prevalence of food insecurity before the COVID-19 pandemic. As a result, strategic policy interventions that include short-term relief and long-term programmatic efforts to support underserved individuals, households, and communities is needed (38).

As well, our research also found that the pandemic has disproportionately affected households with children. While it is estimated that the overall prevalence of food insecurity doubled in the early days of the pandemic, it is estimated that food insecurity among households with children tripled during that time period (37). Again, our analysis showed few differences in our results by sampling strategy, with these differences being limited to comparing convenience and high-risk approaches. Several other studies support our findings, showing consistently that households with children are experiencing high levels of food insecurity during the COVID-19 pandemic (15, 24).

One likely contributor to this trend was the shift to online education, which increased challenges for families that depended on free or reduced price school meals. While federal support such as the Pandemic EBT (P-EBT) program provided additional benefits to families who normally would qualify for these free and reduced-price meals (40), additional hurdles in accessing school meals were reported. For example, the national NFACT survey conducted in the summer of 2020 found that participation in the school meals program dropped during the beginning of the pandemic. Further, between 45 and 55% of survey respondents who utilized school meal pick-ups during the pandemic reported difficulties with availability of delivery, meal pick-up sites being open, and the quantity of food provided (41). Compounding these challenges, low income families with children were more likely to lose income during the COVID-19 pandemic compared to households without children (40). These findings strongly suggest a need for increased support for school food programs, enabling these important programs to ensure that meals reach families in need. Similarly, NFACT sites universally found higher prevalence of food insecurity among households that experienced job or income loss during the pandemic compared to households with no change in employment status, a finding aligned with other recent research (15, 24, 42).

Third, several of our sites conducted repeated surveys, providing insights into the prevalence of food insecurity over time, both within and across regions. Four NFACT study sites have conducted more than one round of surveys (New York City, New York Capital Region, Vermont, and Washington state), while more recent surveys (Massachusetts and the second Washington state survey) were designed to elucidate respondent experiences with food insecurity within the past 30-days, providing a more current understanding of food insecurity prevalence. All follow-up studies found an increasing prevalence of food security as the pandemic continues, with each additional survey demonstrating higher prevalence of food insecurity. All of these rounds of surveys measured food insecurity since the COVID-19 pandemic began, suggesting that additional numbers of individuals continue to become food insecure, even after the initial impacts of COVID-19 have been felt. Likewise, the recent NFACT Massachusetts survey, measuring food insecurity prevalence in the last 30 days, found that nearly 30% of respondents were classified as food insecure at the end of 2020. This prevalence is significantly higher than the pre-pandemic level of 8.4% for this state (13), though it should be noted the pre-pandemic level for Massachusetts respondents was much higher than observed pre-pandemic levels.

This evidence corroborates other studies suggesting that food insecurity levels are likely to persist above pre-pandemic levels for an extended period of time as occurred after the Great Recession and past disasters (13, 17, 20). As noted by Hernandez and Holtzclaw (43), the combined impact of a pandemic and a recession are unique in modern memory. However, the slow recovery from the 2008 Great Recession in the United States is instructive. It took eleven years for food insecurity levels to return to pre-recession levels after the Great Recession; according to national data, food insecurity went from 11.1% in 2007 to 14.6% in 2008, reaching a peak of 14.9% in 2011, and back to 11.1% only in 2018 (13). Similarly, high levels of food insecurity were observed up to five years following other disasters, such as after Hurricanes Katrina and Harvey (20, 44). Besides factors such as age, race/ethnicity, and income, other factors such as support systems, community and generalized self-efficacy are also critical when addressing food insecurity in a post-disaster context. A 5-year follow-up study on Hurricane Katrina revealed that post-disaster food insecurity levels were associated with poor physical and mental health, as well as low social support, generalized self-efficacy, and sense of community (17, 44). Considering that the pandemic has disproportionately affected racial and ethnic minorities, these populations are likely to experience higher levels of food insecurity and be affected by its long-lasting health effects even after the economy recovers. Taken together, these results suggest that the impact of COVID-19 on food security in the U.S. is far from over, and additional support systems and policies will be necessary to continue to alleviate the long-term impacts of the global pandemic and recession.

### Limitations

In presenting our results, we recognize two key limitations. First, research that requires participants to report eating or food-related behaviors is challenged by both recall and social desirability bias (45). Retrospectively asking participants about food insecurity has been shown to lead to overestimation of pre-COVID prevalence of food insecurity (34), suggesting our study may contain similar overestimations. However, we try to address this potential limitation by reporting percent change between pre and during COVID-19 food security, in addition to absolute prevalence of food insecurity, though if the overall pre-pandemic levels of food insecurity are lower, our absolute food insecurity prevalence is likely underestimated. While there has been some skepticism about the high prevalence of food insecurity reported since the COVID-19 pandemic began, our results confirm this high prevalence while providing a more robust measure to benchmark changes. Second, surveys across all research sites included in this study were administered online, limiting respondents to those with computer skills and internet access. This potentially introduced a barrier for some (though not all) elderly or low-income potential respondents (46, 47), as well as those living in rural areas without reliable internet (48). Our study employed a number of methods to overcome this challenge across different sampling strategies. These strategies included partnering with non-profit and community organizations as well as government assistance programs to advertise the survey, and seeking economic representation through sampling targets. Notably, our results show no statistically significant difference between sites using convenience and representative samples, indicating that even for study sites that employed a convenience sampling approach, this potential bias did not have a significant influence on our findings. While differences did emerge when representative and targeted high-risk samples were compared, we argue that this shows the importance of purposeful sampling in target communities.

A number of U.S. studies have explored the impact of COVID-19 on food insecurity prevalence since the beginning of the COVID-19 pandemic, though most have been national samples, modelling efforts, or single site-specific studies. Here, we report the results from a nationwide collaborative effort across 18 study sites and a nationally representative sample, including 22 surveys since the beginning of the COVID-19 pandemic. The scale of our work provides data from more than 27,000 people, and more completely demonstrates the economic hardship the COVID-19 pandemic has had for many people. Consistent increases in food insecurity are prevalent, as well as further evidence that the pandemic has exacerbated racial and ethnic disparities in food insecurity that existed prior to the pandemic. Surveys conducted in study sites more than once also demonstrate an increasing prevalence of food insecurity since the COVID-19 pandemic began, and more recent studies reaffirm that high prevalence of food insecurity, compared to before the COVID-19 pandemic, continues. These findings point to the clear continued need for additional programmatic and policy assistance to provide food insecurity and economic relief. Our future work will continue to conduct additional surveys and comparative analysis to quantify changes in food access, food security, and food assistance use as the U.S. recovers from the COVID-19 pandemic.

## Data Availability

The survey instruments are archived at Harvard Dataverse.

https://dataverse.harvard.edu/dataverse/foodaccessandcoronavirus

## Acknowledgements

This research is conducted as part of The National Food Access and COVID research Team (NFACT), which is implementing common measurements and tools across study sites in the US. NFACT is a national collaboration of researchers committed to rigorous, comparative, and timely food access research during the time of COVID. We do this through collaborative, open access research that prioritizes communication to key decision-makers while building our scientific understanding of food system behaviors and policies.

We are grateful to the Nutrition and Obesity Policy Research and Evaluation Network (NOPREN) for their support of the ad-hoc COVID-19 Food Security Surveys subgroup that shared insights and surveys relevant to this project. We thank Christi Sherlock at The University of Vermont for her assistance with project management.

## Conflicts of Interest

The Authors declare no conflicts of interest.

Funding support:

**Alabama:** No funding to report.

**Arizona**: This project was supported by a COVID-19 seed grant from the College of Health Solutions, Arizona State University.

**California-Bay Area**: This project was supported by the College of Health and Human Sciences, San Jose State University.

**Chicago/Illinois**: This project was supported by the College of Liberal Arts and Social Sciences, DePaul University

**Connecticut**: Generous funding was provided to Foodshare by the Hunger to Health Collaboratory.

**Maine**: We would like to thank the University of Maine School of Food and Agriculture and the George J. Mitchell Center for Sustainability Solutions for their financial support.

**Maryland**: This research was supported by a Directed Research grant from the Johns Hopkins Center for a Livable Future.

**Massachusetts**: Funding for The Greater Boston Food Bank team was provided by the Hunger to Health Collaboratory.

**Michigan**: Funding was provided from Wayne State University faculty startup funds.

National: This research was supported by the College of Health Solutions, Arizona State University with support from the college’s COVID-19 seed grant and the university’s Investigator Research Funds; the University of Arizona College of Agriculture and Life Sciences Rapid COVID-19 seed grant; a Directed Research grant from the Johns Hopkins Center for a Livable Future; and the University of Vermont, the College of Agriculture and Life Sciences, the Gund Institute for Environment, Office of the Vice President of Research, and the UVM ARS Food Systems Research Center.

**New York City:** Special thanks to the Vincentian Institute for Social Action for sponsoring both studies.

**NY State**: Funded by the Natural Hazards Center, Quick Response Grant. The Quick Response program is based on work supported by the National Science Foundation (Award #1635593).

Any opinions, findings, conclusions, or recommendations expressed in this material are those of the author(s) and do not necessarily reflect the views of NSF or the Natural Hazards Center.

**NY Capital Region**: Funding for the Qualtrics Panel Survey was provided by the Foundation for Food and Agriculture Research. Funding for the non-Qualtrics Panel Survey was provided by UAlbany President’s COVID-19 MHD Engaged Researchers Seed Funding Program.

**NY Central/Upstate**: This work was supported by a Cornell Atkinson Center COVID-19 Rapid Response Fund award.

**Utah**: We would like to thank the Utah Create Better Health Program, Utah State University Extension, and the Utah Department of Workforce Services for their support and assistance with this project.

**Vermont**: Funding was provided by The University of Vermont College of Agriculture and Life Sciences and Office of the Vice President of Research, The Gund Institute for Environment, and the UVM ARS Food Systems Research Center.

**Washington**: The WAFOOD survey team wishes to thank the UW Population Health Initiative (UWPHI), the UW School of Public Health (UWSPH), and the Department of Epidemiology for their support.

## State Specific Acknowledgements

**Alabama**: We thank Auburn University at Montgomery, the Alabama Cooperative Extension System at Auburn University, county Supplemental Nutrition Assistance Program Educators, the Alabama Department of Public Health, End Child Hunger in Alabama and the Montgomery Area Food Bank for assisting in dissemination of the survey.

**Arizona**: We thank all the members of the ASU Food policy and environment research group as well as Dr. Aggie J Yellow Horse for their helpful feedback on the survey instrument. We also would like to thank Marina Acosta Ortiz for her assistance with the translation of the survey. This project was supported by a COVID-19 seed grant from the College of Health Solutions, Arizona State University.

**California-Bay Area**: We wish to thank the study participants for their time and dedication in completing the survey. Thank you also to our community partners for helping distribute the survey to potential participants. This project was supported by the College of Health and Human Sciences, San Jose State University.

**Connecticut**: Generous funding was provided to Foodshare by the Hunger to Health Collaboratory.

**Michigan**: Funding was provided from Wayne State University faculty startup funds.

**National**: This research was supported by the College of Health Solutions, Arizona State University with support from the college’s COVID-19 seed grant and the university’s Investigator Research Funds; the University of Arizona College of Agriculture and Life Sciences Rapid COVID-19 seed grant; a Directed Research grant from the Johns Hopkins Center for a Livable Future; and the University of Vermont, the College of Agriculture and Life Sciences, the Gund Institute for Environment, Office of the Vice President of Research, and the UVM ARS Food Systems Research Center.

**New Mexico**: We would like to thank the many community organizations and institutions that assisted with the dissemination of our survey, particularly New Mexico First, New Mexico Thrives, Presbyterian Healthcare Services, and New Mexico State Extension. We also want to thank Gaby Phillips and Aracely Tellez for help with translating the survey into Spanish.

**New York City:** We would like to thank all the New Yorkers, including St. John’s University and its many community partners who participated in this study. Special thanks to the Vincentian Institute for Social Action for sponsoring both studies.

**NY Central**: Our team is very grateful to our partners at the Cornell Cooperative Extension offices in Broome, Cortland, Cayuga, Onondaga, Oswego, and Seneca offices; to the NY Department of Environment and Conservation; and to all the others who helped with survey design, distribution, and applications of findings. We are also grateful to the survey respondents themselves, and doubly so to those who volunteered and who are participating in follow up interviews. This work was supported by a Cornell Atkinson Center COVID-19 Rapid Response Fund award.

**Utah**: We would like to thank the Utah Create Better Health Program, Utah State University Extension, and the Utah Department of Workforce Services for their support and assistance with this project. We would also like to thank the Utah SNAP-eligible individuals who completed the survey.

**Vermont**: We would like to thank many community partners for assisting with the dissemination of the survey especially: Community College of Vermont, Farm to Institution New England, Front Porch Forum, Hunger Free Vermont, Rural Vermont, Salvation Farms, Support and Services at Home (SASH), VT Academy of Nutrition and Dietetics, VT Department of Agriculture, VT Department of Children and Families, VT Department of Health, VT Farm to Plate Network, VT Foodbank, VT Retail and Grocers Association, VT Sustainable Jobs Fund. We would like to thank many community collaborations especially: Funding was provided by The University of Vermont College of Agriculture and Life Sciences and Office of the Vice President of Research, The Gund Institute for Environment, and the UVM ARS Food Systems Research Center.

**Washington**: We wish to thank numerous community partners and stakeholders who helped shape this project. Among those are: WA Department of Health, WA Department of Agriculture, WA Anti-Hunger & Nutrition Coalition, WA SNAP-Ed, KC Local Food Initiative, Northwest Harvest, Washington State University (WSU) Extension, United Way of WA, and numerous food banks, food pantries, charitable organizations community organizations, county health departments, and local health jurisdictions.

## Author Contributions

The research design, data collection, and analysis of individual site data was conducted by site-level NFACT teams (see supplementary materials for all authors). A.W.B, L.A.C., M.M.D., M.T.N., G.A.P., M.S.R., S.R. R.S., and R.Z. wrote and edited the paper. M.T.N. conducted the analysis comparing the study site types and food security prevalence. M.T.N. had primary responsibility for final content. All authors read and approved the final manuscript.

