## Supplementary Materials for "A Multi-Site Analysis of the Prevalence of Food Security in the United States, before and during the COVID-19 Pandemic"

Full Author List

Our manuscript represents a comprehensive group of study sites and surveys, which necessitates a large author list given the scale of the research. The first nine authors are the core writing team, who, in addition to data design, collection and analysis for their or multiple sites, also wrote and edited the paper. These authors are listed in the manuscript submission, and bolded in the table below. Please note that with the exception of the first author (M.T. Niles), the remaining core authors are listed alphabetically on purpose. In addition, our NFACT team consists of a larger group of authors who contributed to data design, collection and analysis for their sites. These individuals are listed in alphabetical order immediately following the core author list.

| **Person** | **Study Site** | **Affiliation** | **Institution** |
| --- | --- | --- | --- |
| **Meredith T. Niles** | Vermont, National | Department of Nutrition and Food Sciences, Gund Institute for Environment | University of Vermont |
| **Alyssa W. Beavers** | Michigan | Department of Nutrition and Food Science | Wayne State University |
| **Lauren A. Clay** | New York | Health Administration & Public Health Department | D'Youville College |
| **Marcelle M. Dougan** | California | Department of Public Health and Recreation | San José State University |
| **Giselle A. Pignotti** | California | Department of Nutrition, Food Science, and Packaging | San José State University |
| **Stephanie Rogus** | New Mexico | Department of Family and Consumer Sciences | New Mexico State University |
| **Mateja R. Savoie-Roskos** | Utah | Department of Nutrition, Dietetics and Food Sciences | Utah State University |
| **Rachel E. Schattman** | Maine | School of Food and Agriculture | University of Maine, Orono |
| **Rachel M. Zack** | Massachusetts | Business and Data Analytics | The Greater Boston Food Bank |
| Francesco Acciai | Arizona | College of Health Solutions | Arizona State University |
| Deanne Allegro | Alabama | Department of Kinesiology | Auburn University at Montgomery |
| Emily H. Belarmino | Vermont, National | Department of Nutrition and Food Sciences, Gund Institute for Environment | University of Vermont |
| Farryl Bertmann | Vermont, National | Department of Nutrition and Food Sciences | University of Vermont |
| Erin Biehl | National, Maryland | Johns Hopkins Center for a Livable Future | Johns Hopkins University |
| Nick Birk | Massachusetts | Business and Data Analytics | The Greater Boston Food Bank |
| Jessica Bishop-Royse | Chicago | Faculty Scholarship Collaborative, College of Liberal Arts and Social Sciences | DePaul University |
| Christine Bozlak | New York Capital Region | Health Policy, Management, and Behavior | University at Albany- State University of New York |
| Brianna Bradley | Maryland | Johns Hopkins Bloomberg School of Public Health | Johns Hopkins University |
| Barrett P. Brenton | New York City | Center for Civic Engagement | Binghamton University |
| James Buszkiewicz | Washington | Department of Epidemiology | University of Washington |
| Brittney N. Cavaliere | Connecticut | Institute for Hunger Research & Solutions | Connecticut Food Bank/Foodshare |
| Young Cho | Wisconsin | Joseph J Zilber School of Public Health | University of Wisconsin-Milwaukee |
| Eric M. Clark | Vermont | Department of Plant and Soil Science | University of Vermont |
| Kathryn Coakley | New Mexico | Department of Individual, Family, and Community Education | University of New Mexico |
| Jeanne Coffin-Schmitt | New York - Central/Upstate | Department of Natural Resources | Cornell University |
| Sarah M. Collier | Washington | Department of Environmental and Occupational Health Sciences | University of Washington |
| Casey Coombs | Utah | Department of Nutrition, Dietetics and Food Sciences | Utah State University |
| Anne Dressel | Wisconsin | College of Nursing | University of Wisconsin-Milwaukee |
| Adam Drewnowski | Washington | Department of Epidemiology | University of Washington |
| Tom Evans | Arizona | School of Geography, Development and Environment | University of Arizona |
| Beth J Feingold | New York Capital Region | Department of Environmental Health Sciences | University at Albany- State University of New York |
| Lauren Fiechtner | Massachusetts | Department of Gastroenterology and Nutrition | MassGeneral Hospital for Children |
| Kathryn J. Fiorella | New York -Central/Upstate | Department of Population Medicine and Diagnostic Sciences and Master of Public Health Program | Cornell University |
| Katie Funderburk | Alabama | Alabama Cooperative Extension System | Auburn University |
| Preety Gadhoke | New York City | Department of Pharmacy Administration and Public Health, College of Pharmacy and Health Sciences | St. John's University (at the time of study administration) |
| Diana Gonzales-Pacheco | New Mexico | Department of Individual, Family, and Community Education | University of New Mexico |
| Amelia Greiner Safi | New York - Upstate | Department of Population Medicine and Diagnostic Sciences and Master of Public Health Program: Department of Communication | Cornell University |
| Sen Gu | New York CIty | Department of Pharmacy Administration and Public Health, College of Pharmacy and Health Sciences | St. John's University |
| Karla L. Hanson | New York - Central/Upstate | Department of Population Medicine and Diagnostic Sciences and Master of Public Health Program | Cornell University |
| Amy Harley | Wisconsin | Joseph J Zilber School of Public Health | University of Wisconsin-Milwaukee |
| Kaitlyn Harper | Maryland, National | Department of International Health, Bloomberg School of Public Health | Johns Hopkins University |
| Akiko S. Hosler | New York Capital Region | Department of Epidemiology and Biostatistics | University at Albany- State University of New York |
| Alan Ismach | Washington | Department of Environmental and Occupational Health Sciences | University of Washington |
| Anna Josephson | Arizona, National | Department of Agricultural and Resource Economics | University of Arizona |
| Linnea Laestadius | Wisconsin | Joseph J Zilber School of Public Health | University of Wisconsin-Milwaukee |
| Heidi LeBlanc | Utah | Department of Nutrition, Dietetics and Food Sciences | Utah State University |
| Laura R. Lewis | Washington | Community and Economic Development | Washington State University |
| Michelle M Litton | Michigan | Department of Nutrition and Food Science | Wayne State University |
| Katie S. Martin | Connecticut | Institute for Hunger Research & Solutions | Connecticut Food Bank/Foodshare |
| Shadai Martin | New Mexico | Department of Family and Consumer Sciences | New Mexico State University |
| Sarah Martinelli | Arizona | College of Health Solutions | Arizona State University |
| John Mazzeo | Chicago/Illinois | Master of Public Health Program, College of Liberal Arts and Social Sciences | DePaul University |
| Scott C. Merrill | Vermont | Department of Plant and Soil Science, Gund Institute for Environment | University of Vermont |
| Roni Neff | Maryland, National, New York Capital Region | Department of Environmental Health & Engineering, Bloomberg School of Public Health; Johns Hopkins Center for a Livable Future | Johns Hopkins University |
| Esther Nguyen | Washington | Center for Public Health Nutrition | University of Washington |
| Punam Ohri-Vachaspati | Arizona, National | College of Health Solutions | Arizona State University |
| Abigail Orbe | Connecticut | Institute for Hunger Research & Solutions | Connecticut Food Bank/Foodshare |
| Jennifer J. Otten | Washington | Department of Environmental and Occupational Health Sciences | University of Washington |
| Sondra Parmer | Alabama | Alabama Cooperative Extension System | Auburn University |
| Salome Pemberton | New York City |  | Hunter College, City University of New York |
| Zain Al Abdeen Qusair | Chicago/Illinois | Master of Public Health Program, College of Liberal Arts and Social Sciences | DePaul University |
| Victoria Rivkina | Chicago/Illinois | Master of Public Health Program, College of Liberal Arts and Social Sciences | DePaul University |
| Joelle Robinson | Maryland, National | Department of Health, Behavior and Society, Bloomberg School of Public Health | Johns Hopkins University |
| Chelsea M. Rose | Washington | Department of Epidemiology | University of Washington |
| Saloumeh Sadeghzadeh | New York City | School of Management | Binghamton University |
| Brinda Sivaramakrishnan | Washington | Health, Business, & Professional Services | Tacoma Community College |
| Mariana Torres Arroyo | New York Capital Region | Department of Environmental Health Sciences | University at Albany- State University of New York |
| McKenna Voorhees | Utah | Department of Nutrition, Dietetics and Food Sciences | Utah State University |
| Kathryn Yerxa | Maine | Cooperative Extension | University of Maine, Orono |
